# Changes in premature birth rates during the Danish nationwide COVID-19 lockdown: a nationwide register-based prevalence proportion study

**DOI:** 10.1101/2020.05.22.20109793

**Authors:** Gitte Hedermann, Paula L Hedley, Marie Bækvad-Hansen, Henrik Hjalgrim, Klaus Rostgaard, Porntiva Poorisrisak, Morten Breindahl, Mads Melbye, David M Hougaard, Michael Christiansen, Ulrik Lausten-Thomsen

## Abstract

**Objectives:** To explore the impact of COVID-19 lockdown on premature birth rates in Denmark

**Design:** Nationwide register-based prevalence proportion study.

**Participants:** 31,180 live singleton infants born in Denmark between March 12, and April 14, during 2015 to 2020

**Main outcome measures:** Main outcome measure was the odds ratio of premature birth, per preterm category, during the lockdown period compared with the calendar match period in the five previous years.

**Results:** A total of 31180 newborns were included in the study period, of these 58 were born extremely premature (gestational age below 28 weeks). The distribution of gestational ages was significantly different (p = 0·004) during the lockdown period compared to the previous five years. The extremely premature birth rate during the lockdown was significantly lower than the corresponding mean rate for the same dates in the previous years (odds ratio 0·09 [95 % CI 0·01 − 0·04], p < 0·001). No significant difference between the lockdown and previous years was found for other gestational age categories.

**Conclusions:** The birth rate of extremely premature infants decreased significantly (~90 % reduction) during the Danish nationwide lockdown from a stable rate in the preceding five years. The reasons for this decrease are unclear. Identification of possible causal mechanisms might stimulate changes in clinical practice. Ideally, some cases of extreme prematurity are preventable which may decrease infant morbidity and mortality.

## Introduction

The World Health Organization (WHO) declared the new coronavirus disease, COVID-19, a Public Health Emergency of International Concern on the January 30, 2020.^1^ Subsequently, on the basis of more than 20 000 confirmed cases and almost 1 000 deaths in Europe, a pandemic was declared on March 12, 2020.^2^ This led to an almost global lockdown that has been a crucial element in slowing the spread of the virus.^3^ Despite the start and scale of the lockdown varying between countries, the lockdown has literally quieted the planet as even seismic noise has been reduced due to changes in human activity.^4^ Beyond controlling transmission of the virus, the consequences of the global lockdown have been widespread including economic disruptions,^5^ and environmental impacts.^6 7^ Additionally, the lockdown has affected virtually all branches of medicine and brought about changes in patterns of hospital contacts for conditions other than COVID-19.^8^

The consequences of SARS-CoV-2 infections in pregnancy and early infancy are only just being reported, but maternal-foetal transmission appears to be rare^9^ and the majority of SARS-CoV-2-infected pregnancies do not develop major complications.^10^ Although perinatal death has been reported^10^, most SARS-CoV-2 positive neonates appear to be only mildly affected.^10^

Prematurity is a complex and challenging pathophysiological condition associated with increased risk of long term morbidity and mortality^11^ and it is the leading cause of death in children under five years of age.^12 13^ Global prematurity rates are approximately 10 %, but vary from 4·5 % in some European countries to 15-18 % in some parts of Africa and Asia.^12 14^ During the pandemic child birth will continue, and some children will inevitably be born prematurely. The aetiology of premature birth and preterm labour is multifaceted and linked to a wide range of socio-demographic, medical, obstetric, foetal, psychosocial, and environmental factors.^15^ Still, it is only partly understood and approximately two-thirds of premature births occur without an evident risk factor.^15^

In Denmark, a nationwide lockdown was declared from March 12,2020.^16^ Effective from that date, childcare facilities, schools and universities were closed, all non-essential public servants were sent home, private employers were urged to ensure that as many people as possible worked from home, gatherings of more than ten people were prohibited, and the borders were closed to foreign visitors.^16^ Epidemiological reports from the Danish authorities show that the lockdown resulted in a flattening of the epidemic curve.^17^ The number of deaths per day and number of hospitalized patients due to COVID-19 peaked around April 1, 2020.^17^ As of May 20, 2020, 554 have died from COVID-19 in Denmark equivalent to 96 deaths per million inhabitants.^18^ Agradual lifting of lockdown restrictions began on April 15, 2020 as schools and childcare facilities started reopening.^19^

Anecdotal observations from neonatal intensive care units suggest that there were fewer extremely premature births during the lockdown period. It is likely that the lockdown itself - with its changes in work environment, social interactions, and focus on hygiene – has reduced exposure to infectious agents as well as impacted premature birth rates.

To elucidate if any association exists between nationwide lockdown and premature births, we assessed the distribution of gestational age among all live born singleton births in Denmark during the most rigorous part of the nationwide lockdown and compared it with corresponding distributions in previous years.

## Methods

### Study Design and Data Sources

The study was a nationwide prevalence proportion study with premature births as cases, term pregnancies as controls, and birth during the lockdown period as exposure. In Denmark, all newborns have been offered a centralised screening for an expanding number of congenital conditions since 1975. Clinical data on the births have been stored together with the collected and analysed dried blood spots samples (DBSS) at Statens Serum Institut in the Danish National Screening Biobank (DNSB) since 1982.^20^ The participation is nearly complete as judged nationally as well as locally,^21^ and the need to receive and analyse the DBSS within three days after birth makes the DNSB an updated source of information on Danish pregnancies. We used the DNSB to identify children born in Denmark during the most rigorous part of the lockdown period (March 12, to April 14, 2020)^16 19^ and during the corresponding calendar period in the previous five years (2015 to 2019). We also identified births in the period January 20, to February 22, for all years 2015 – 2020 (n= 32 070).

To limit the influence of other determinants of timing of birth, we considered only singletons, for whom information on date of birth and gestational age at delivery, was retrieved. The DNSB database records data on gestational age (reported in completed weeks) at delivery as reported by the midwives based on the information from the hospital charts. We categorised gestational age at birth as extremely premature (before 28 weeks); very premature (28^+0^ weeks – 31^+6^ weeks); moderate/late premature (32^+0^ weeks – 36^+6^ weeks); term (37^+0^ weeks – 41^+6^ weeks); and late term (after 42^+0^ weeks).

### Statistical Analysis

Likelihood-ratio based tests, estimates and confidence intervals regarding changes in composition of gestational age at birth categories between the lockdown period and the consolidated reference period for 2015-2019 were obtained from a series of logistic regressions. Kaplan-Meier curves and frequency plots were used to illustrate variations in gestational ages between the birth cohorts studied, differences between gestational age categories for the periods under study were evaluated by log-rank tests. Statistical analyses were run in SAS (v9.4) and R (v3.6.1).

### Data and Ethics Approvals

Statens Serum Institut has approval from the Danish Data Protection Agency (DPA) to conduct register-based studies and the current study was approved by the DPA officer, approval no: 20/04753 at Statens Serum Institut. Studies based solely on register data do not require further ethics committee approval as per Danish laws and regulations.

### Patient involvement

There were no funds or time available to include patient and public involvement in this study. However, we have an urgent requirement to disseminate the results of the research publicly and have measures in place to do so.

## Results

We included a total of 31,180 live singleton infants born in Denmark from March 12, to April 14, during 2015 to 2020. Births were distributed into gestational age categories as shown in Table 1. The total number of singleton births during lockdown in 2020 (n = 5 162) did not differ statistically significantly from the other years (mean births per year (March 20 – April 14): 5 203·6, SD ± 221·4; p=0·24). We identified 1·566 premature births (gestational age below 37 weeks) in total from singleton pregnancies (5·02 %).

**Table 1:** Gestational age categories and the distribution of singleton births throughout the study periods. (March 12, - April 14, (2015-2020)).

|  | Gestational age (GA)<br>(Weeks + days) | N | Percent | mean GA | SD |
| --- | --- | --- | --- | --- | --- |
| **Extremely preterm** | ≤ 27+6 | 58 | 0·19 | 25·7 | 1·31 |
| **Very preterm** | 28+0 - 31+6 | 177 | 0·57 | 29·7 | 1·07 |
| **Moderate/late preterm** | 32+0 - 36+6 | 1 331 | 4·27 | 34·9 | 1·26 |
| **Term** | 37+0 - 41+6 | 28 947 | 92·84 | 39·6 | 1·15 |
| **Late Term** | ≥ 42+0 | 667 | 2·14 | 42·0 | 0·16 |
| **Total** | all births | 31 180 | 100 | 39·4 | 1·81 |

Logistic regression analyses demonstrated that the distribution of gestational age in 2020 differed highly significantly from the previous years (p = 0·004). The proportion of extremely and very premature births (gestational age below 32 weeks) (shown in Figure 1) was significantly different between the 2020 nationwide lockdown and the same calendar period from the previous five years (p = 0·003). However, the difference was solely due to a reduction in extremely premature to 0·19/1000 births during the 2020 nationwide lockdown compared with an average of 2·19/1000 births for the five previous years (p < 0·001) (Table 2, Figure 1). In order to compare the distribution of gestational age categories across a time period not influenced by the lockdown, we retrieved data on births from January 20, to February 22, in 2020 and the preceding five years (n = 32 070). It is evident that the reduction in proportion of extremely premature births was not present in the months immediately prior to the lockdown (Figure 1 inset). The Kaplan-Meier curves depicting births as a function of gestational age (Figure 2) indicates a significant shift in gestational age at birth among extremely and very premature (gestational age < 32 weeks) births during the lockdown period (p= 0·01), this shift is not noted in later weeks (gestational age < 32 weeks, p = 0·8).

**Figure 1:**
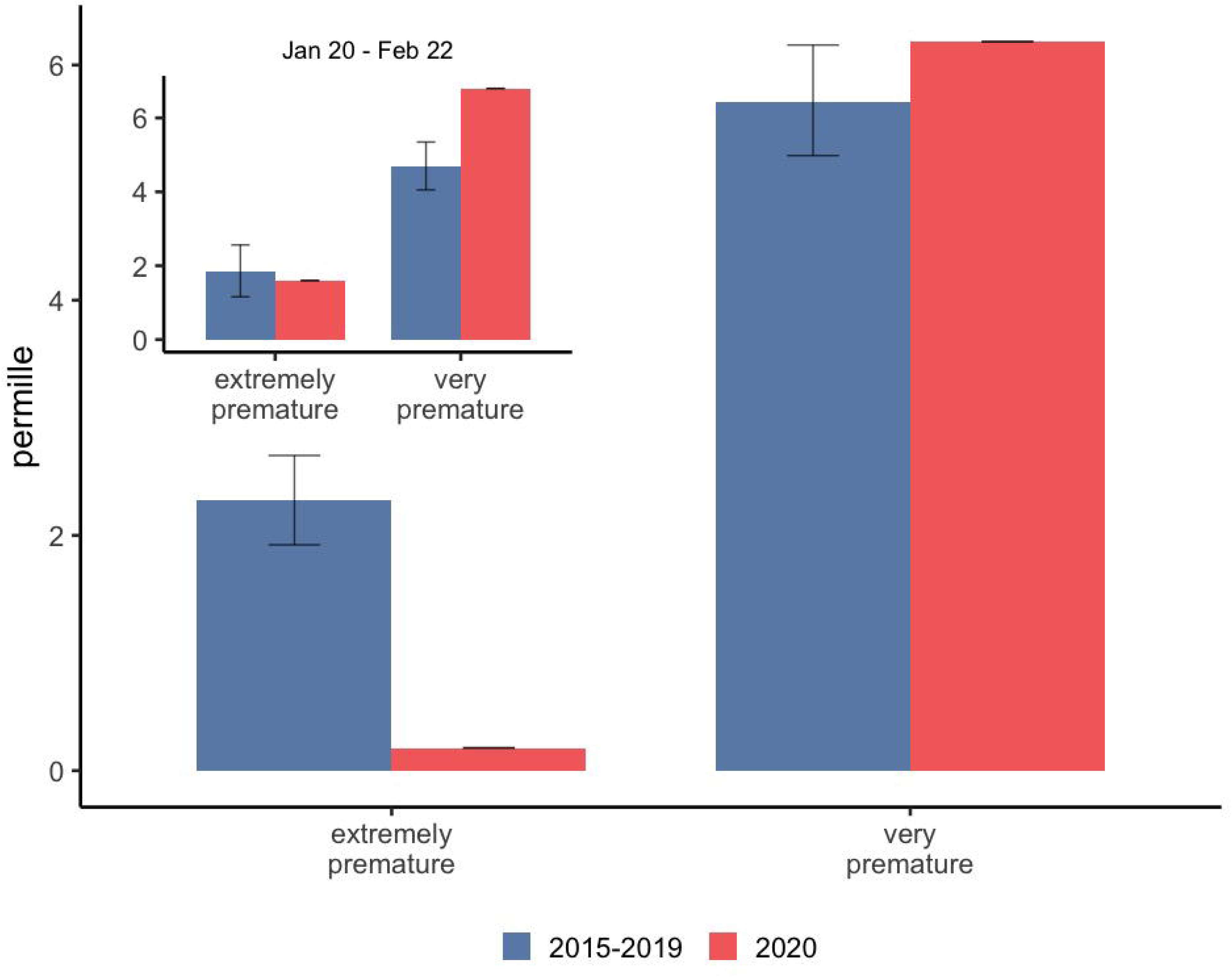
The proportion of extremely premature and very premature births (permille of all births in the time period) during the lockdown period (March 12 – April 14, 2020) compared with aggregated birth data for the previous five years during the same date range (March 12 – April 14, 2015-2019). **Inset graph:** A comparison of extremely premature and very premature births born between January 20, and February 22, 2020 and an aggregate from that date range for the previous five years (January 20 to February 22, 2015-2019). Extremely premature (before 28^+0^ weeks’ gestation) and very premature (28+^0^ – 31+^6^ weeks’ gestation).

**Figure 2:**
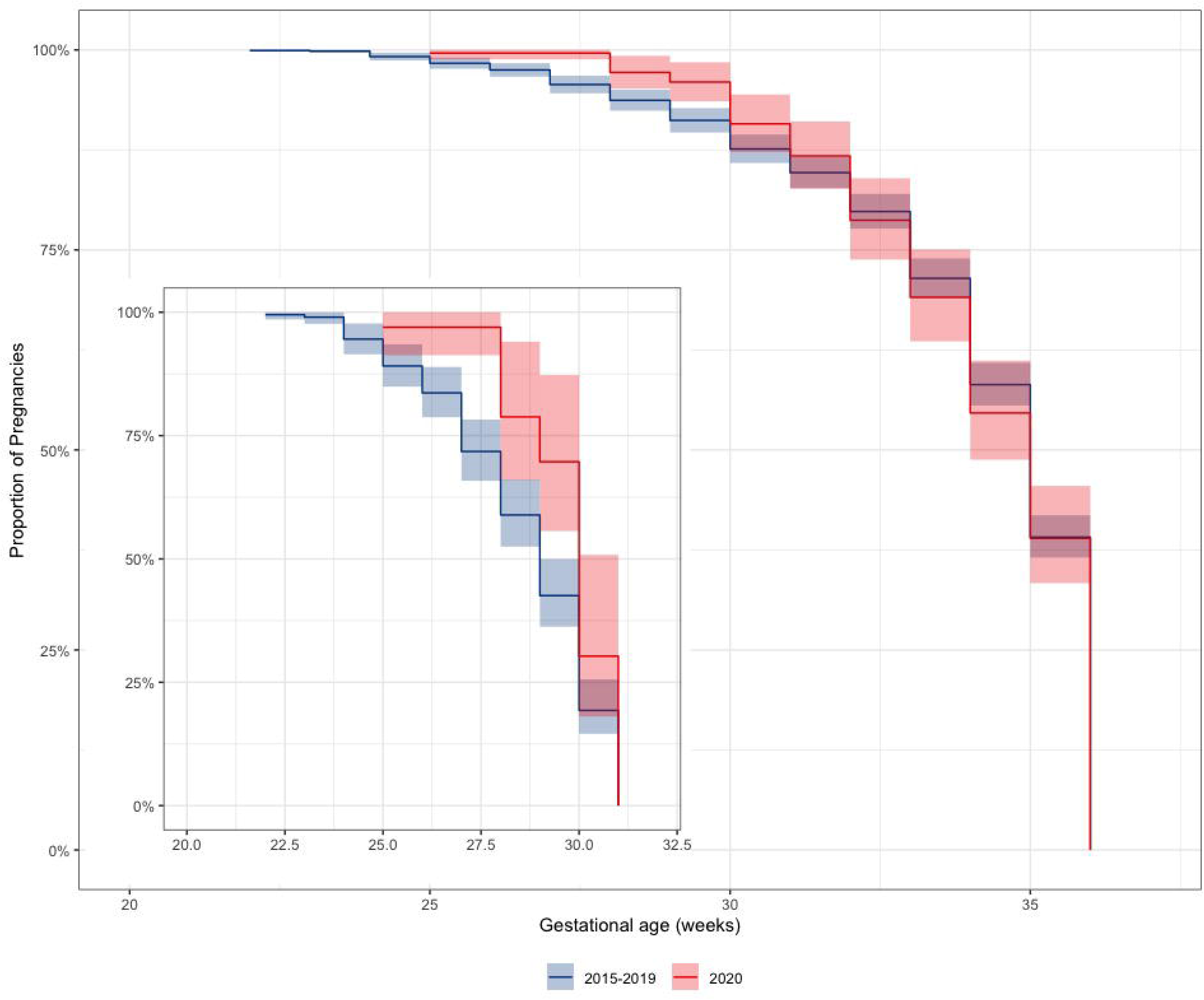
Kaplan-Meier curve comparing gestational age at birth for gestational ages <37 weeks for the lockdown period (March 12 – April 14, 2020) and the aggregate of the same date range (March 12 – April 14) for the previous five years (2015-2019). **Inset graph:** indicates the Kaplan-Meier curve for gestational ages < 32 weeks.

**Table 2:** The distribution of births permille, by gestation age category for the lockdown period (March 12 – April 14, 2020) compared with consolidated data from March 12 – April 14, 2015-2019.

|  | Prevalence (per mille) |  | OR | 95 % CI |  | p-value |
| --- | --- | --- | --- | --- | --- | --- |
|  | 2015-2019 | 2020 |  |  |  |  |
| **Extremely preterm** | 2·19 | 0·19 | 0·09 | 0·01 | 0·40 | <0·001 |
| **Very preterm** | 5·57 | 6·20 | 1·11 | 0·75 | 1·61 | 0·589 |
| **Moderate/late preterm** | 42·85 | 41·84 | 0·98 | 0·84 | 1·13 | 0·742 |
| **Term** | 927·70 | 931·81 | 1·06 | 0·95 | 1·20 | 0·293 |
| **Late term** | 21·68 | 19·95 | 0·92 | 0·74 | 1·13 | 0·430 |

## Discussion

In this study, we analysed nationwide data on live births from the DNSB and identified a marked decrease in the number of extremely premature births. This is the first study to report a potential effect of a nationwide lockdown on extremely premature birth rates. Although too early to draw any definitive conclusions, we believe that these findings and their potential implications merits to be signalled to the public immediately. Despite modern neonatal intensive care, the complexity of prematurity makes it the leading cause of death in neonates and children under five. Therefore, any prevention of preterm labour is a key factor in reducing perinatal and early paediatric morbidity and mortality.

A premature birth may be initiated by multiple factors, but in most cases the precise mechanism cannot be identified.^22^ The COVID-19 lockdown has drastically changed our lives by reducing physical interactions, increasing our focus on hygiene, changing our working environment, and lowering air pollution. This unusual situation is likely to have influenced several risk factors for premature birth.

### Potential lockdown effects on modifiable risk factors of premature birth

Several risk factors for premature birth are known to give rise to increased systemic maternal inflammation^23^, which along with other immunologically mediated processes are believed to play a part in the preterm birth syndrome.^24^ It is possible that the increased focus on hygiene, strict physical distancing, and home confinement during the lockdown period have influenced the overall inflammatory state of pregnant women. Thus, we have already seen reports documenting a significant lowering of the incidence of influenza and other viral and bacterial infections as a result of the COVID-19 lockdown.^23^

The literature on a potential link between work and premature birth is contradictory,^24 25^ yet the changes in physical work and activity caused by the lockdown restrictions may equally play a role in the observed reduction of premature births. Additionally, the decrease in air pollution may play a role as air pollution, particularly the anthropogenic PM_2.5_, has been estimated in meta-analyses to be associated with 18 % of premature births globally.^26^ The Centre for Research on Energy and Clean Air has estimated a potential reduction in premature birth in Europe due to reduced air pollution.^27^ Therefore, the lockdown associated improved air quality could also be a contributing factor to the observed reduction in the number of extremely premature births.

### Premature birth during lockdown

We found no significant differences in the rates of the very premature, moderate premature, term or post term births, which may reflect that no such differences exist, or that the differences are too subtle to be detected. However, it is noteworthy that we observed a non-significant but slightly increased number of very premature births. It is possible that whatever impact the lockdown had on risk factors for premature birth, it served to simply postpone extremely preterm labour in some high-risk pregnancies, although this impact was not sufficient to avoid premature births altogether. For extremely premature infants the chance of survival increases dramatically with increasing gestational age at delivery. Identification of risk factors that may result in postponement of time of delivery for such infants may therefore have large implications for their chance of survival.

### Strengths and limitations

Our study has several strengths. As Denmark has excellent registers with a very high coverage,^28^ we believe the data accurately reflects the current prematurity rates in Denmark. It is based on reliable mandatorily reported data from the entire country. Because exposure (the lockdown) is independent of the recorded outcome, differential misclassification is not considered to be an issue. Although it is possible that a larger number of pregnancies resulted in intrauterine death and that these pregnancies were classified as late abortions, this seems unlikely to explain our observations as it collides with reports from obstetric departments.^29^

Importantly, this study is observational and the association between the decreased number of extremely premature births and nationwide lockdown is not necessarily causal. As such, this data needs to be confirmed in other countries, although international discrepancies regarding changes in premature birth rates could reflect the variation in baseline premature birth rates as well as differences in implementation of national lockdowns around the world. Future studies should also aim to elucidate potential causalities.

### Conclusions

Our data indicates that the occurrence of extreme prematurity may be further reduced through preventive measures. If this tendency is confirmed, future studies may even identify causal mechanisms that may be applicable outside a lockdown. Possibly, lessons learned during the COVID-19 pandemic and lockdown may contribute to improved guidelines leading to fewer extremely premature births and thus decrease infant morbidity and mortality.

What is already known on this topic

- Prematurity, particularly extreme prematurity, defined as gestational age below 28 weeks, has a high morbidity, and is considered the primary cause of mortality in children under five years old.
- Global overall prematurity rates are approximately 10 %, but large regional variation exists.
- The aetiology of preterm labour and premature birth is multifaceted and linked to a wide range of socio-demographic, medical, obstetric, foetal, psychosocial, and environmental factors.

What this study adds

- The rates of premature birth decreased during the COVID-19 lockdown and it is possible that elements of the lockdown (e.g. generally reduced infection load caused by increased focus on hygiene, physical distancing, reduced work and physical activity, and improved quality of air) are beneficial for reducing extreme prematurity and potentially reducing infant mortality.

## Data Availability

Data sharing: The process of accessing data from the Danish National Biobanks is detailed here https://www.danishnationalbiobank.com/access. No additional data are available.

## Acknowledgements

This research was conducted using the Danish Neonatal Screening Biobank and the Danish National Biobank resource, funded by the Novo Nordisk Foundation.

## Footnotes

**Contributors:** GH*, PLH*, DMH, MC# and ULT# designed the study. GH, MBH collected the data. PLH*, KR, MC#, and ULT# performed statistical analyses. GH*, PLH*, MC# and ULT# co-wrote first draft. All authors contributed to the interpretation of the data and critically revised the manuscript. All authors had full access to tables and figures in the study and can take responsibility for the integrity of the data and the accuracy of the data analysis. ULT# and MC# share the responsibility of guarantors. The corresponding authors (ULT, MC) attests that all listed authors meet authorship criteria and that no others meeting the criteria have been omitted.

* contributed equally as first authors, # contributed equally as corresponding authors

**License for publication:** The corresponding author has the right to grant on behalf of all authors and does grant on behalf of all authors, a worldwide licence to the Publishers and its licensees in perpetuity, in all forms, formats and media (whether known now or created in the future), to i) publish, reproduce, distribute, display and store the Contribution, ii) translate the Contribution into other languages, create adaptations, reprints, include within collections and create summaries, extracts and/or, abstracts of the Contribution, iii) create any other derivative work(s) based on the Contribution, iv) to exploit all subsidiary rights in the Contribution, v) the inclusion of electronic links from the Contribution to third party material where-ever it may be located; and, vi) licence any third party to do any or all of the above.

**Competing interests:** All authors have completed the ICMJE uniform disclosure form at www.icmje.org/coi_disclosure.pdf and declare: no support from any organisation for the submitted work; MB has a patent (NeoHelp) with royalties paid, all other authors reported no financial relationships with any organisations that might have an interest in the submitted work in the previous three years; no other relationships or activities that could appear to have influenced the submitted work.

**Data sharing:** The process of accessing data from the Danish National Biobanks is detailed here https://www.danishnationalbiobank.com/access. No additional data are available.

**Transparency:** The corresponding author (ULT) affirms that the manuscript is an honest, accurate, and transparent account of the study being reported; that no important aspects of the study have been omitted; and that any discrepancies are disclosed.

